# Combining a prioritization strategy and functional studies nominates 5’UTR variants underlying inherited retinal disease

**DOI:** 10.1101/2023.06.19.23291376

**Authors:** Alfredo Dueñas Rey, Marta del Pozo Valero, Manon Bouckaert, Filip Van Den Broeck, Malena Daich Varela, Mattias Van Heetvelde, Marieke De Bruyne, Stijn Van de Sompele, Miriam Bauwens, Jamie Ellingford, Hanne Lenaerts, Quinten Mahieu, Dragana Josifova, Genomics England Research Consortium, Carlo Rivolta, Andrew Webster, Gavin Arno, Carmen Ayuso, Julie De Zaeytijd, Bart P. Leroy, Elfride De Baere, Frauke Coppieters

## Abstract

**Background:** 5’ untranslated regions (5’UTRs) are essential modulators of protein translation. Predicting the impact of 5’UTR variants is challenging and typically not performed in routine diagnostics. Here, we present a combined approach of a comprehensive prioritization strategy and subsequent functional assays to evaluate 5’UTR variation in two large cohorts of patients with inherited retinal diseases (IRDs).

**Methods:** We performed an isoform-level re-analysis of retinal RNA-seq data to identify the protein-coding transcripts of 378 IRD genes with highest expression in retina. We evaluated the coverage of these 5’UTRs by different whole exome sequencing (WES) capture kits. The selected 5’UTRs were analyzed in whole genome sequencing (WGS) and WES data from IRD sub-cohorts from the 100,000 Genomes Project (n = 2,417 WGS) and an in-house database (n = 1,682 WES), respectively. Identified variants were annotated for 5’UTR-relevant features and classified into 7 distinct categories based on their predicted functional consequence. We developed a variant prioritization strategy by integrating population frequency, specific criteria for each category, and family and phenotypic data. A selection of candidate variants underwent functional validation using diverse experimental approaches.

**Results:** Isoform-level re-quantification of retinal gene expression revealed 76 IRD genes with a non-canonical retina-enriched isoform, of which 20 display a fully distinct 5’UTR compared to that of their canonical isoform. Depending on the probe-design 3-20% of IRD genes have 5’UTRs fully captured by WES. After analyzing these regions in both IRD cohorts we prioritized 11 (likely) pathogenic variants in 10 genes (*ARL3*, *MERTK*, *NDP*, *NMNAT1*, *NPHP4*, *PAX6*, *PRPF31*, *PRPF4*, *RDH12*, *RD3*), of which 8 were novel. Functional analyses further supported the pathogenicity of 2 variants. The *MERTK*:c.-125G>A variant, overlapping a transcriptional start site, was shown to significantly reduce both luciferase mRNA levels and activity. The *RDH12*:c.-123C>T variant was found *in cis* with the reported hypomorphic *RDH12*:c.701G>A (p.Arg234His) variant in 11 patients. This 5’UTR variant, predicted to introduce an upstream open reading frame, was shown to result in reduced RDH12 protein but unaltered mRNA levels.

**Conclusions:** This study demonstrates the importance of 5’UTR variants implicated in IRDs and provides a systematic approach for 5’UTR annotation and validation that is applicable to other inherited diseases.

## Background

Among the numerous non-coding regulatory regions found across the human genome, 5′ untranslated regions (5’UTRs) are major determinants of post-transcriptional control and translation efficiency^1, 2^. These regions, with an average length of about 200 nucleotides in humans, are located immediately upstream from protein-coding sequences and include the Kozak consensus sequence around the AUG start codon^3^. 5’UTRs harbor numerous *cis*-regulatory elements such as internal ribosomal entry sites (IRES) and upstream open reading frames (uORFs), which can recruit scanning ribosomes and initiate translation^4^. In particular, uORFs, defined by an upstream start codon in-frame with a stop codon preceding the end of the primary open reading frame, can decrease downstream protein expression up to 80%^5^. Furthermore, 5’UTRs can serve as platforms for the formation of secondary and tertiary mRNA structures like stem-loops, hairpins, and RNA G-quadruplexes which further influence mRNA translation^6,7^. Given the critical importance of 5’UTRs as modulators of protein expression, genetic variation within these regions can contribute to disease pathogenesis, as shown by several examples in a wide range of inherited diseases^8–11^.

With the expanding implementation of massively parallel sequencing technologies in clinical practice, the rare disease field has witnessed a true paradigm shift enabling improved molecular diagnosis for many affected individuals^12–15^. Despite these advancements, and although this varies greatly across individuals with diverse clinical indications, a significant fraction of cases with suspected Mendelian disorders remains unsolved^12,13, 16–18^.

To date, 5’UTR variants are often not evaluated because of the multiple challenges they entail. Firstly, detection relies on inclusion of these non-coding regions in targeted resequencing panels or whole genome sequencing, which has been limited to date^19^. Secondly, interpretation of the effect is more difficult due to different mechanisms^8^ and our relative lack of understanding of the functional consequences of 5’UTR variants. Finally, functional evidence is necessary to support their pathogenicity and hence confirm the molecular diagnosis. Recently, to close the annotation and interpretation gap, several *in silico* tools^20–26^ and guidelines^27^ have been developed, although their specific applicability to the accurate and comprehensive interpretation of the diverse pathogenic mechanism of 5’UTR variants is yet to be fully established.

A clear instance of both the effectiveness and remaining challenges of clinical large-scale genome analyses can be found in inherited retinal diseases (IRDs)^28–31^. IRDs comprise a genetically and phenotypically diverse constellation of visually debilitating conditions affecting over 2 million people globally^32,33^. With over 290 known disease genes^34^, diverse modes of inheritance^29^, intersecting phenotypes^35,36^, and available automated approaches to variant interpretation, establishing a genetic diagnosis in IRD patients can be challenging. Even after the use of unbiased approaches such as whole exome (WES) and whole genome sequencing (WGS), up to 50% of cases remain unsolved ^15,28, 37–39^, and hence without the possibility of potential clinical intervention including gene therapy-based treatments^40,41^.

Recent studies have revealed an important contribution of non-coding variation to IRDs, particularly with the identification of deep-intronic mis-splicing variants^39,42, 43–50,51^. Moreover, there is a growing body of evidence supporting the emerging role of genetic variation affecting *cis*-regulatory regions in the molecular pathogenesis of IRDs^45,48, 52–56^. However, thus far only a few 5’UTR variants have been reported to be implicated in ocular diseases, including IRDs^48,53,54, 57–59^, but no large-scale studies have been performed yet.

In this work, we conducted a systematic evaluation of 5’UTR variation in IRD genes using WGS and WES data derived from two IRD cohorts comprising 2,417 participants of the 100,000 Genomes Project^12^ and 1,682 local cases, respectively. Firstly, we obtained a comprehensive selection of 5’UTRs of the most abundant canonical as well as non-canonical protein-coding IRD gene isoforms by performing transcript-level re-quantification of retinal expression data. We then screened these regions for variants in both IRD cohorts and developed a prioritization strategy to identify candidate pathogenic 5’UTR variants. This allowed us to identify 11 potentially causative 5’UTR variants, 10 of which were found in unsolved cases. Functional validation of the predicted pathogenetic mechanism was performed for 4 of these, further supporting the potential implication in disease for 2 variants. Overall, we show that 5’UTRs represent understudied targets of non-coding variation that can provide novel molecular diagnoses in IRDs and demonstrate the importance of reassessing these regions in existing exome and genome sequencing data.

## Methods

### Re-analysis of retinal RNA-seq data and selection of retina-enriched isoforms of IRD genes

We retrieved paired-end FASTQ files (GSE115828) derived from human postmortem retina samples characterized by Ratnapriya *et al.,* (2019)^60^. Only samples derived from donor retinas showing no features of age-related macular degeneration were evaluated (n=102). Transcripts were quantified through pseudoalignment by *Kallisto*^61^ (v.0.46.1) using default parameters. Abundance estimates in transcripts per million (TPM) for all annotated transcripts (*Ensembl* human release 107) were retrieved. A custom R (v.4.0.2) script was used for annotating each transcript with its corresponding gene and biotype and flagging them as canonical and/or belonging to the Matched Annotation from NCBI and EMBL-EBI (MANE) and MANE Plus Clinical set (version 1.0)^62^. For each group of isoforms belonging to each gene, we computed the average of abundance estimates across all samples and the isoform exhibiting the highest average among its corresponding group was deemed retina-enriched. This dataset was further filtered by retaining only protein-coding isoforms derived from selected genes, namely IRD genes listed in either the *Retinal disorders panel* (v2.195) from Genomics England PanelApp^63^ or RetNet (https://sph.uth.edu/retnet/) (**Table S1**). Additionally, we integrated cap analysis gene expression sequencing (CAGE-seq) data derived from fetal and adult retina (FANTOM5^64^ Robust Peak Set using an expression RLE normalized threshold >1) to evaluate the confidence of the annotated transcription start sites (TSS) of the selected transcripts in retina.

To aid tiering of variants, the Genomics England PanelApp^63^ gene color-coded rating system was used: genes with diagnostic-grade rating, borderline evidence and research candidates were flagged with green, amber, and red, respectively. To further assess the retina-enrichment of specific isoforms, their loci were inspected using an integration of multiple publicly available multi-omics datasets derived from human retina (**Table S2**).

### Comparison of 5’UTRs of canonical and non-canonical isoforms and selection of 5’UTR variant search space

The exact genomic coordinates of the start and end positions of the 5’UTRs of all genes were downloaded from *Ensembl biomart* (Human Ensembl Genes 107, GRCh38.p13) and filtered to only include the transcripts defined above. We first assessed the number of exons composing these 5’UTRs to identify which IRD genes have spliced 5’UTRs. For each IRD gene for which a retina-enriched non-canonical isoform was identified, we compared the canonical and non-canonical 5’UTRs and computed their respective overlap. The calculated overlaps were then used to classify these genes into three different categories, namely genes with transcripts displaying: (i) fully distinct 5’UTRs, (ii) partly overlapping 5’UTRs, and (iii) fully overlapping 5’UTRs.

For each 5’UTR exon of all selected transcripts, we defined a near-splice region by including 25-bp intronic sequence up- and downstream of its respective splice donor and acceptor sites (excluding promoter and coding sequences). The resulting coordinates were annotated with their corresponding transcript identifier and gene name and stored into a sorted BED file (herein after referred to as *5’UTR analysis file*) for downstream variant assessment.

### Evaluation of 5’UTR capture by whole-exome sequencing

We assessed the performance of exome captures of the selected 5’UTRs based on the designs provided by the kits which were mostly used for the generation of our in-house WES data, namely the SureSelect Human All Exon V6 and SureSelect Human All Exon V7 (Agilent Technologies), as well as a selection of the most recent versions of commonly used kits from 4 different providers: SureSelectXT Human All Exon V8 (Agilent Technologies), KAPA HyperExome V2 (Roche), Twist Exome 2.0 (Twist), Illumina Exome Panel v1.2 (Illumina). BED files containing the genomic coordinates of the capture regions were downloaded from the corresponding design catalogs and intersected with the coordinates of the 5’UTRs of interest using *bedtools intersect* (v2.26.0)^65^ with default parameters. For uniformity, only the capture regions were used to compare between the different designs. Additionally, for the SureSelect Human All Exon V6 and SureSelect Human All Exon V7 kits (Agilent Technologies) both the strict union of all regions covered by baits and a version padded by ±50bp extending into intronic regions were considered for the intersections. A custom Python (v.3.6.8) script was then used to compute for each IRD gene the length proportion (%) of its 5’UTR captured by these kits.

### Search of 5’UTR variants in IRD genes submitted to ClinVar

The ClinVar database (ClinVar) was downloaded in a tab-delimited format^66^ directly from the FTP site (https://ftp.ncbi.nlm.nih.gov/pub/clinvar/; version from 2023-03-18) and pre-filtered to keep only entries from the GRCh38.p13 build. The resulting file was further filtered to retrieve variants located within the regions defined in the *5’UTR analysis file.* Large copy number gain/loss variants extending into coding regions as well as variants with a protein-altering or synonymous annotation for the canonical transcript were removed from this analysis.

### Cohort selection

To assess the contribution of 5’UTR genomic variation to IRDs, individuals from two different cohorts were selected for this study: (i) 2,397 participants (2,100 probands) from a sub-cohort of the Rare Disease arm of the 100,000 Genomes Project (Genomics England –GE– cohort) affected by posterior segment abnormalities (**Table S3**); (ii) an IRD sub-cohort of 1,682 cases (1,030 probands) with a WES analysis performed at the Center for Medical Genetics Ghent (CMGG cohort). In both cases, sequencing data aligned to GRCh38 build were included.

The 100,000 Genomes Project Protocol has ethical approval from the HRA Committee East of England – Cambridge South (REC Ref 14/EE/1112). This study was registered with Genomics England within the *Hearing and sight* domain under Research Registry Projects 465. This study was approved by the ethics committee for Ghent University Hospital (B6702021000312) and performed in accordance with the tenets of the Helsinki Declaration and subsequent reviews.

### Sequencing and variant analysis

We interrogated WGS and WES data from the two cohorts described above to detect germline single-nucleotide variants (SNVs) and small insertions and deletions (indels) overlapping the regions defined in the *5’UTR analysis file*. The sequencing and bioinformatic pipelines used for processing genome data derived from the participants of the GE cohort have been described previously^12^.

Samples from the CMGG cohort were tested using WES with the SureSelect Human All Exon V6, SureSelect Human All Exon V7 (Agilent Technologies) or HyperExome (Roche) enrichment kits and sequenced on HiSeq 3000 or NovaSeq 6000 instruments (paired-end 150 cycles) (Illumina). Reads were aligned to the human reference genome (GRCh38 build) with *BWA* (v0.7.15)^67^ and *GATK HaplotypeCaller* (v3.82)^68^ was used for calling SNVs and indels. Resulting variant call format (VCF) files from both cohorts were subsequently parsed based on the regions of interest using *BCFtools* (v1.9)^69^. Only variants satisfying the filter criteria for sequencing depth (DP>10) and genotype quality (GQ>15) were retrieved.

### Variant annotation and prioritization

From each cohort, a file containing unique variants was created and formatted so that it could be annotated with the *Ensembl Variant Effect Predictor*^70^ (VEP, release 107). Apart from gene and transcript information, each variant was annotated with frequency data retrieved from gnomAD^71^ (genomes, v3.1.2) allele frequencies, splicing (dbscSNV^72^, SpliceAI^23^, MaxEntScan^73^), pathogenicity predictions (EVE^74^, CADD^75^), and regulatory data^76^. The recently developed *UTRannotator*^20^ tool was used to annotate variants that create or disrupt uORFs. The classical^77^ and retinal^78^ Kozak consensus sequences were considered altered when variants were identified within the position range −1 to −10 relative to the main AUG (canonical start codon). In addition, nucleotide frequency plots corresponding to the Kozak sequences of the retina-enriched and not retina-enriched transcripts evaluated in this study were generated using were generated with *WebLogo*^79^ (v.3.7.12). Annotation data tables containing predicted changes in translational efficiency^80^ and secondary structure minimum free energy affecting double-stranded RNA or G4 quadruplex structures^81^ were downloaded (*5utr [’suter’]*: https://github.com/leklab/5utr) and queried for the variant positions using *tabix*^82^ (v.1.7-2); further evaluation of changes in mRNA secondary structure was performed using the *Ufold*^83^ (v.1.2), *REDfold*^84^ (v1.14.alpha), and *MXfold2*^85^ (v0.1.2) tools. To assess whether variants were located within TSS relevant to retinal gene expression, we made use of the CAGE-seq data described above. Translation initiation-related feature data from the Human Internal Ribosome Entry Sites (IRES) Atlas^86^ was also queried to annotate variants found within these regions involved in cap-independent translation initiation.

Annotated 5’UTR variants were classified into the following 7 categories: (i) uAUG gained, (ii) change in existing uORF, (iii) alteration of classical or retinal primary Kozak context, (iv) splicing, (v) change in translational efficiency (TE), (vi) change in secondary structure minimum free energy, and (vii) overlapping a retinal transcription start site and/or an IRES. Variants were first selected when their minor allele frequency was lower than 2% in all populations. The resulting variants were further filtered using specific criteria for each category: (i) uAUG created in a strong or moderate Kozak context, (ii) natural uAUG loss, (iii) variant located in positions −3, −4, −5, −6, or −9, (iv) any SpliceAI Delta Score (DS_AG, DS_AL, DS_DG, DS_DL) higher than 0.2, (v) |log2FC|≥0.5, (vi) |FC| ≥1.5, (vii) variant located within a TSS or IRES. Variants for which the inheritance pattern and phenotype of the patient were compatible with the gene in which the 5’UTR variant was identified were selected as candidates. This prioritization procedure is depicted in **Figure 1**. For comparing the proportion of variants for each category between cohorts, the statistical analysis was performed in R using the χ^2^-test. Finally, for each candidate variant identified in unsolved cases, an additional screening was performed to discard (likely) pathogenic variants, both SNVs and structural variants (SVs), in other IRD genes that could provide an alternative molecular diagnosis. For the cases from the GE cohort in which the identified 5’UTR variant remained as candidate, we requested to have a clinical collaboration with Genomics England.

**Figure 1.**
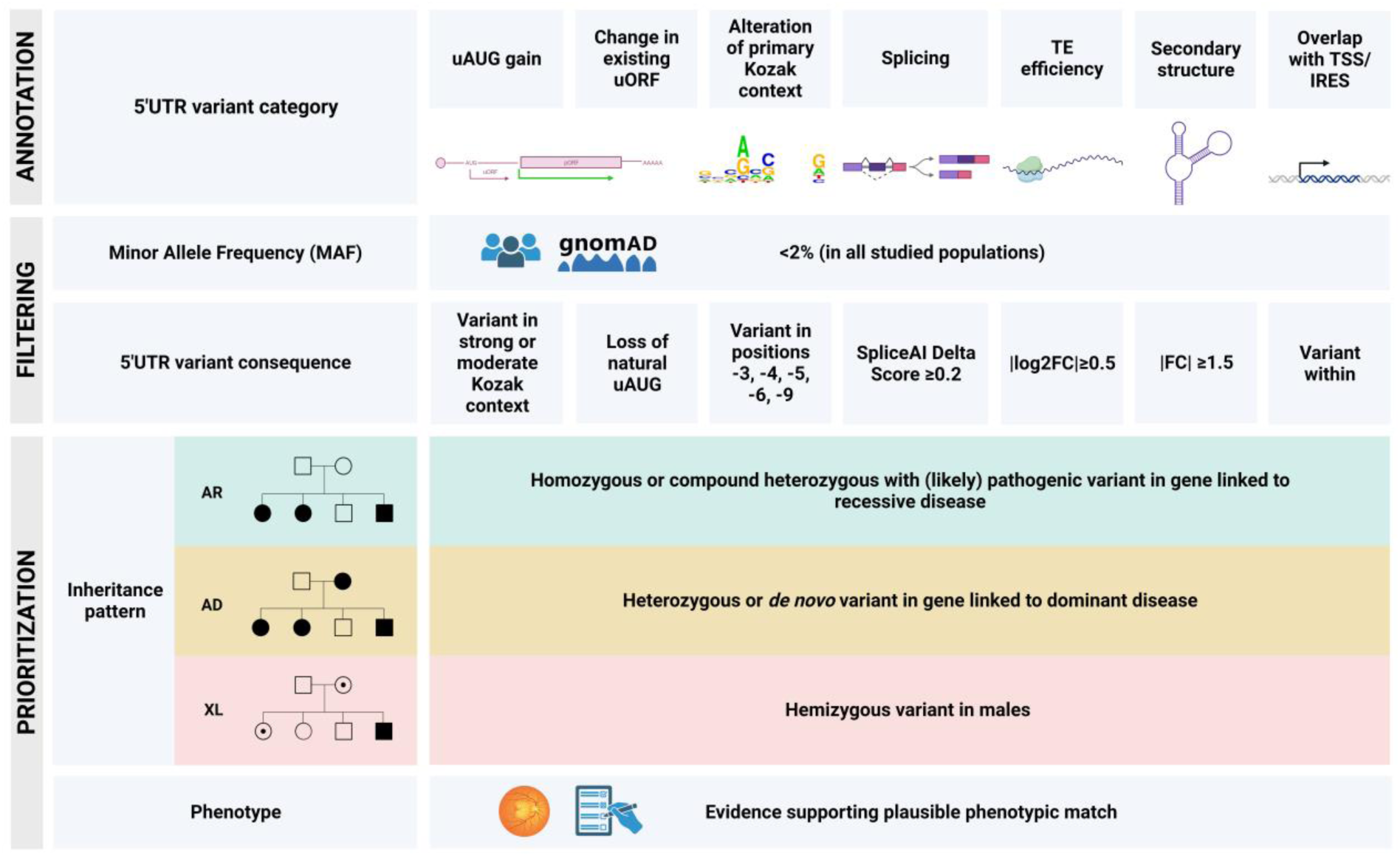
Functional annotation of 5’UTR variants, filtering, and prioritization strategy followed in this study. A combination of *in silico* tools (*see Methods*) was used to annotate 5’UTR variants, which were then classified into 7 functional categories. For each of these categories, specific criteria were established for prioritizing variants with a more likely functional impact (*bottom*). Only variants with a minor allele frequency (MAF) <2% were further studied. The following information was then reviewed: inheritance pattern of the family (AR including sporadic cases; AD; XL) and clinical features. For the selection of candidate variants, both had to be in agreement with the reported mode of inheritance and phenotype associated with the gene in which the 5’UTR variant was found. Abbreviations: AD: autosomal dominant; AR: autosomal recessive; FC: fold change; IRES: internal ribosomal entry site; TE: translational efficiency; TSS: transcription start site; uAUG: upstream AUG; uORF: upstream open reading frame.

### Cell culture

ARPE-19 (ATCC, CRL-2302™) and HEK-293T cells (ATCC CRL-3216™) cells were grown in either Dulbecco’s minimal essential medium (DMEM) with phenol red (Thermo Scientific Life Technologies) or DMEM:F12 (Gibco) medium supplemented with 10% fetal bovine serum (Gibco), 1% penicillin-streptomycin (Gibco), 1% non-essential amino acid solution (Gibco), and 0.1% amphotericin B (Gibco), respectively. Cells were cultured at 37°C and 5% CO2 and tested for mycoplasma contamination prior to use.

To perform functional studies of the identified 5’UTR *ARL3* variant, lymphocytes from affected carriers (n=2) were isolated from EDTA blood using Lymphoprep (STEMCELL technologies). For each sample two cultures were started in RPMI medium with 10% fetal bovine serum and substituted with interleukin-2 and phytohaemagglutinin. One of both cultures was treated for four hours with puromycin (200 µg/mL) prior to RNA extraction to suppress nonsense-mediated decay.

### Cloning and mutagenesis

In order to evaluate the functional effect of 3 selected 5’UTRs variants (*RDH12*:c.-123C>T, *MERTK*:c.-125G>A, *PAX6*:c.-44C>T), we cloned the wild-type 5’UTR of interest (IDT gBlock) into a psiCHECK™-2 dual luciferase vector (Promega) using the Cold Fusion cloning kit (Sanbio BV). The recombinant vectors were then amplified in One Shot TOP10 Chemically Competent *E. coli* cells (Invitrogen) and purified using the NucleoBond Xtra Midi kit (Filter Service S.A). For the generation of the overexpression construct to further assess the *RDH12*:c.-123C>T variant, we designed a gBlocks™ fragment comprising the wild-type 5’UTR and coding sequence (CDS) of *RDH12*; both sequences were modified to include downstream Myc and FLAG in-frame tags to evaluate the translation of the uORF introduced by the *RDH12*:c.-123C>T variant and RDH12 protein levels, respectively. These fragments were then cloned into a pcDNA™3.1^(+)^ (Invitrogen) vector by restriction-ligation cloning and the recombinant vectors were amplified and purified as described above. For all constructs, 5’UTRs variants were introduced using the Q5 Site-Directed Mutagenesis Kit (NEB) using variant-specific primers designed with the *NEBaseChanger* tool. The sequence of each insert was confirmed by Sanger sequencing using the BigDye Terminator v3.1 kit (Life Technologies) and/or long-read whole plasmid sequencing (Plasmidsaurus). A schematic overview of these constructs is shown in **Figure S1**. All primer sequences can be found in **Table S4**.

### Dual luciferase assays

ARPE-19 cells were seeded in a 24-well plate (Greiner Bio-One BVBA) at a density of 50,000 cells/well in 1mL of medium without antibiotics to reach 70-90% confluence at transfection (18-24h after plating). Cells were transfected at a 3:1 reagent to plasmid DNA ratio using the TransIT-X2® Dynamic Delivery System (Mirus Bio) according to the manufacturer’s instructions. After 24 h, cells were lysed, and luciferase activity was detected using the Dual-Glo® Luciferase Assay System (Promega) in a Glomax 96-Microplate Luminometer (Promega). Each transfection was performed in triplicate and each experiment was repeated at least three times to ensure reproducibility. For each well, the ratio of *Renilla* luciferase activity was normalized to *Firefly* luciferase activity. A custom R script was used to evaluate the effect of each variant on luciferase activity through a linear mixed effects model (implemented in the *lme4* package^87^) having set the luciferase vector as fixed effect and the biological replicate as random effect.

### RNA isolation and quantitative polymerase chain reaction (qPCR)

Total RNA was extracted using either the RNeasy Mini kit® (Qiagen) (ARPE-19 and HEK-293T cells) or the Maxwell RSC simply RNA kit (Promega) (cultured lymphocytes) according to the manufacturer’s instructions. Isolated RNA underwent DNase treatment (ArcticZymes, Tromsø, Norway) prior to cDNA synthesis with the iScript cDNA Synthesis Kit (Bio-Rad Laboratories). For each cDNA sample, qPCR assays were prepared using SsoAdvanced Universal SYBR Green Supermix (Bio-Rad Laboratories) and run on LightCycler 480 System (Roche). Data were analyzed with *qbase+*^88^ (CellCarta, v.3.4) and normalized either to a set of housekeeping genes (*YWHAZ*, *HPRT1*, *HMBS*, *SDHA*) or *Firefly* luciferase for normalization of *Renilla* luciferase mRNA levels. All primer sequences can be found in **Table S4**. Statistical analyses were performed in R using the Wilcoxon rank sum test.

### Overexpression and immunoblotting

To perform immunoblotting of RDH12 and the predicted peptide encoded by the uORF introduced by the *RDH12*:c.-123C>T variant, ARPE-19 cells were seeded in 12-well plates (Greiner Bio-One BVBA) at a density of 100,000 cells/well, allowed to settle overnight, and then transfected with the wild-type and mutant *RDH12* overexpression vectors using the TransIT-X2® Dynamic Delivery System (Mirus Bio) according to the manufacturer’s instructions. The pcDNA™3.1^(+)^ (Invitrogen) backbone vector was transfected as negative control. Four hours prior to RNA and protein isolation, cells were treated with 10µM MG-132 proteasome inhibitor (Merck Life Science). For total protein extraction, cells were lysed with RIPA Buffer (Sigma-Aldrich) including protease inhibitory cocktail (Roche Diagnostics), phosphatase inhibitory cocktail 2, and phosphatase inhibitory cocktail 3 (Sigma-Aldrich). Protein concentrations were measured using the Pierce^TM^ BCA Protein Assay kit (Fisher Scientific). After centrifugation and reduction with 1M DTT (Sigma-Aldrich), protein lysates were subjected to sodium dodecyl sulfate-polyacrylamide gel electrophoresis using either NuPAGE™ 4–12% Bis-Tris (RDH12 blot) or Novex™ 16% Tricine (uORF blot) Protein Gels (Fisher Scientific) with a ladder (Precision Plus Protein All Blue Standards, Bio-Rad Laboratories). Proteins were then transferred to a nitrocellulose membrane using the iBlot 2 Dry Blotting System (Thermo Fisher Scientific). Membranes were blocked for 2 hours at room temperature in 2% ECL™ Blocking Agent (Cytiva Amersham) and incubated at 4°C overnight with anti-FLAG (1:1000, F1804, Merck Life Science) or anti-Myc (1:1000, ab9106, Abcam) primary antibodies. Membranes were subsequently incubated for 2 hours at room temperature with the appropriate horseradish-peroxidase-conjugated secondary antibody (1:2500, 7076S or 7074S, Cell Signaling Technologies) and revealed with the SuperSignal™ West Dura Extended Duration Substrate (Fisher Scientific). Membranes were scanned with an Amersham Imager 680 system (GE Healthcare Life Sciences). Protein quantification was performed by firstly stripping the membranes with Restore™ PLUS Western Blot Stripping Buffer (Thermo Scientific), incubating them for 1 hour at room temperature with a primary antibody against β-tubulin (1:2500, ab6046, Abcam) and for 2 hours with a horseradish-peroxidase-conjugated secondary antibody (1:2500, 7074S, Cell Signaling Technologies). RDH12 (FLAG) signal intensity quantification was achieved using *ImageJ* (NIH, v.1.50i) and normalized to the amount of β- tubulin.

To further assess the translation of the peptide encoded by the uORF introduced by the *RDH12*:c.-123C>T variant, transfected HEK-293T cells were lysed as previously described, and protein lysates incubated at 4°C overnight with Dynabeads Protein A (Thermo Fisher Scientific), to which the anti-Myc primary antibody (1:1000, ab9106, Abcam) was bound. Bound proteins were subsequently eluted and subjected to anti-Myc Western Blot analysis as described above. Statistical analyses were performed in R using the Student’s t-test.

### Screening of *RDH12*: c.-123C>T variant

The *RDH12*:c.-123C>T 5’UTR variant was shared with members of the European Retinal Disease Consortium (ERDC: https://www.erdc.info/) to evaluate its pathogenicity and identify additional carriers of the *RDH12*:c.701G>A (p.Arg234His) missense variant to confirm its *cis*-configuration. Primer sequences for PCR and Sanger sequencing are listed in **Table S4**.

## Results

### Isoform-level re-quantification of retinal gene expression reveals differential 5’UTRs of IRD genes

To obtain a relevant selection of 5’UTRs for downstream variant screening, we performed a transcript-level re-analysis of RNA-seq data to identify protein-coding non-canonical isoforms with relevant retinal expression i.e., higher than its respective canonical isoform, to be retained in addition to the canonical isoforms. A total of 454 canonical and non-canonical transcripts belonging to 378 IRD genes were thus selected (with maximum of 2 isoforms per gene) (**Table S5**), from which their 5’UTRs were retrieved, resulting in 638 genomic regions (**Table S6**). Considering these 454 transcripts, the 5’UTRs of approximately 62% of IRD genes (233/378) are only part of the first coding exon while 33% of IRD genes (126/378) have exclusively transcripts with spliced 5’UTRs, i.e., their 5’UTR comprise additional full non-coding exons. A minor fraction (19/378) of genes has isoforms with both spliced and non-spliced 5’UTRs (**Figure 2A**; **Table S7**). Of the 76 IRD genes for which a retina-enriched non-canonical isoform was identified, 20 display a fully distinct 5’UTR compared to the one of their corresponding canonical isoforms (**Figure 2B**; **Table S8**). For six of these, the non-canonical TSS is further supported by CAGE-seq data derived from both adult and fetal retina (**Table S5**; **Figure S2**). Two remarkable examples are *CRB1* and *RIMS2*, for which the non-canonical isoforms (ENST00000681519.1: 48.38±14.97 TPM; ENST00000436393.6: 41.58±12.35 TPM)were found to be more abundant in retina compared to their respective canonical isoforms (ENST00000367400.8: 2.54±3.88 TPM; ENST00000696799.1: 12.19±3.85 TPM). These findings were also supported by the integration of multiple multi-omics datasets derived from human retina (**Figure S3; Table S2**).

**Figure 2.**
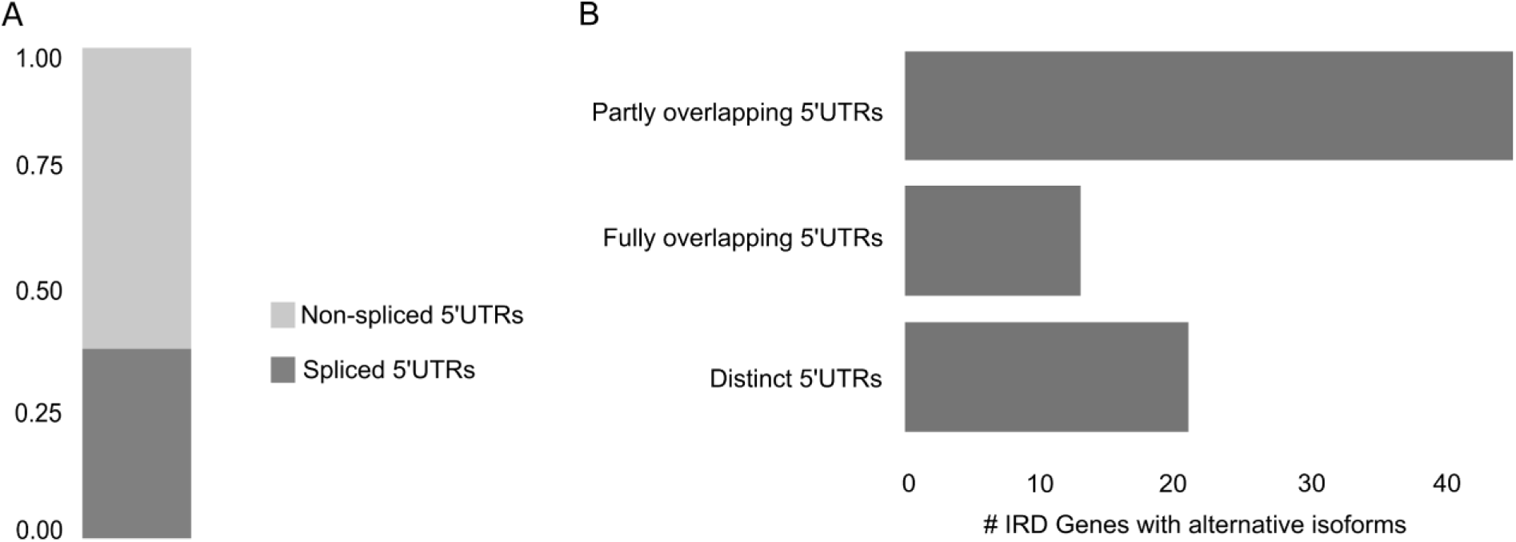
Characterization of 5’UTRs of IRD genes. **(A)** Representation of the proportion of IRD gene isoforms based on the structure of their 5’UTRs. **(B)** Classification of IRD genes with a retina-enriched isoform differing to the canonical one based on the comparison of their respective 5’UTRs.

### A substantial fraction of 5’UTRs can be captured by whole-exome sequencing

In view of the large volume of existing WES data, we evaluated the performance of six recent and commonly used commercial exome capture designs from four different providers on the selected 5’UTRs of IRD genes. The kits considered in this analysis were found to display a variable performance with an average coverage of the selected 5’UTRs ranging from 7% (Twist Exome 2.0) to 39% (Illumina Exome Panel v1.2) (**Figure 3A**; **Table S9**). Besides, the fraction of IRD genes with 5’UTRs fully captured ranged from 3% (10/378) to 20% (73/378). Regarding the kits that were mostly used in-house (SureSelect Human All Exon V6 and SureSelect Human All Exon V7), although a slightly higher 5’UTR capture was observed for the kit with a higher amount of probes (SureSelect Human All Exon V6) (**Figure S4**; **Table S10**), we found that approximately 15% of IRD genes (57/378) have the selected 5’UTRs fully captured. This fraction increased up to 39% (148/378) (**Figure 3B**; **Table S10**) if a padded bait design, including all regions that can be confidently genotyped, was considered.

**Figure 3.**
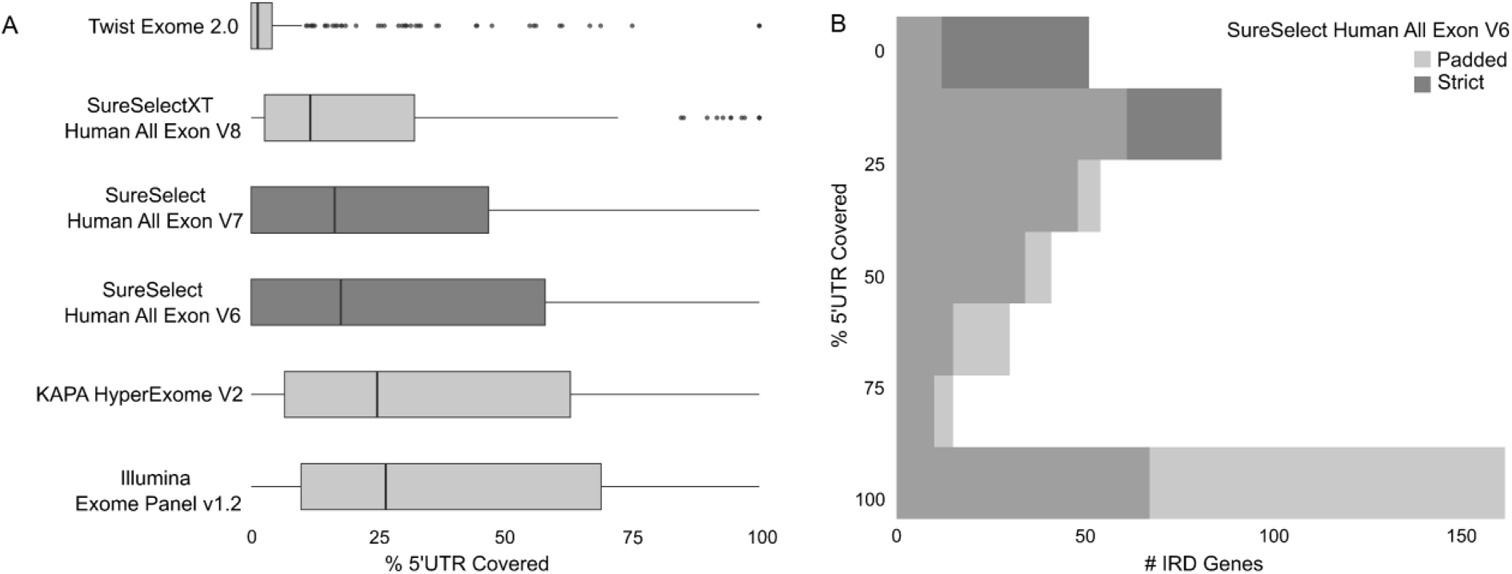
Evaluation of 5’UTR capture by whole-exome sequencing (WES). **(A)** Boxplots showing the capture performance (*x-axis*) of commonly used commercial exome capture designs on the selected 5’UTRs. The kits that were mostly used for the generation of our in-house WES data are highlighted in *darker gray*. **(B)** Histogram representing the portions of 5’UTRs (*y-axis*) of the selected IRD genes which are captured by the SureSelect Human All Exon V6 kit (Agilent Technologies) considering a *strict* or a *padded* design (*see Methods*).

### A retrospective analysis of 5’UTR variants reported in IRD genes reveals a majority classified as Variants of Uncertain Significance (VUS)

A total of 1,547 5’UTR variants in IRD genes have been submitted to the ClinVar database thus far (**Table S11**). All variants except 4 have been clinically interpreted. Only 2% (31/1,547) of the variants have been classified as pathogenic or likely pathogenic, of which 32% (10/31) and 16% (5/31) were found in the *PAX6* and *NMNAT1* genes, respectively. On the other hand, 34% (527/1,547) of the 5’UTR variants have been classified as benign or likely benign, of which no single gene was found to account for more than 5% of these variants. While 5% (81/1,547) of the variants had conflicting interpretations of pathogenicity, the largest fraction comprising 58% (904/1,547) of all variants have been classified as VUS.

### Analysis of 5’UTRs in WGS and WES data of two IRD cohorts reveals rare and ultra-rare variants predicted to affect 5’UTR function

To systematically assess the contribution of 5’UTR genomic variation to IRDs, we performed a variant analysis within the selected 643 genomic regions in affected individuals from two different IRD sub-cohorts, namely the GE (n=2,397 WGS) and CMGG (n=1,682 WES) cohorts. We identified a total of 2,898 and 381 distinct 5’UTR variants within the GE and CMGG cohorts, respectively. The majority of these variants (2,637/2,898 and 334/381) had a minor allele frequency (MAF) lower than 2% in all populations and a substantial fraction (506/2,898 and 92/381) was found to be absent from all reference population public databases. To aid the interpretation of these 5’UTR variants, we classified them into 7 (non-mutually exclusive) categories according to their *in silico* predicted functional consequences (**Figure 1**). A summarized overview of the number of variants that remained after classification and category-specific filtering can be found in **Table 1**. Out of the 1,450 remaining variants (**Dataset S1**), 370 were present in more than one category. Of the remaining variants assigned to a single category, the bulk corresponded to variants overlapping a retinal TSS or an IRES (761/1,450; 221 -TSS-, 488 -IRES-, 52 -TSS & IRES-), followed by variants with a predicted change in secondary structure minimum free energy (204/1,450). When comparing the number of variants in each category between the two cohorts, we observed statistically significant differences in the number of variants overlapping a retinal TSS and/or IRES, for which the proportion of variants was higher for the WGS-based GE cohort compared to the WES-based CMGG cohort (~55% and ~35% respectively, p<0.05). These differences are most likely due to the expanded capture of regions including TSS allowed by the use of WGS in the GE cohort. Additionally, it is noteworthy that for the IRD genes with 5’UTRs harboring IRES (92/378), these elements were found to span on average 61% of the 5’UTR and even its entire length for certain genes (12/92) (**Table S12**).

**Table 1.** Summarized overview of the number of 5’UTR variants that remained after functional classification and filtering.

|  | GE cohort | CMGG cohort | Total | % |
| --- | --- | --- | --- | --- |
| uAUG gained | 30 | 5 | 35 | 2.41 |
| Change in existing uORF | 17 | 1 | 18 | 1.24 |
| Alteration of classical or retinal primary Kozak context | 16 | 6 | 22 | 1.52 |
| Splicing | 29 | 10 | 39 | 2.69 |
| Change in translational efficiency | 1 | 0 | 1 | 0.07 |
| Change in secondary structure minimum free energy | 194 | 10 | 204 | 14.07 |
| Overlapping a retinal transcription start site or an IRES | 692 | 69 | 761 | 52.48 |
| > 1 category | 276 | 94 | 370 | 25.52 |
| **Total** | **1255** | **195** | **1450** |  |

Abbreviations: CMGG, Center for Medical Genetics Ghent; GE, Genomics England; IRES, internal ribosomal entry site; uORF, upstream open reading frame.

### Systematic variant evaluation allows the prioritization of candidate pathogenic 5’UTR variants

To enhance the likelihood of identifying potential pathogenic 5’UTR variants, we prioritized the ones with higher predicted impact based on specific criteria for each category (**Figure 1**) and for which the inheritance patterns and reported phenotypes fitted the genes in which the 5’UTR variants had been identified. The application of this prioritization procedure is illustrated in **Table S13**. We prioritized 11 candidate pathogenic 5’UTR variants. Of these, 10 were found in unsolved cases and 8 have not been reported before. Of note, although initially identified through WGS, 6 of these variants would also be covered by WES. An overview of the genetic, phenotypic, and *in silico* prediction details of these variants is shown in **Table 2**. The pedigrees of these families including variant segregation are shown in **Figure S5**.

**Table 2.** Summarized overview of the 5’UTR variants that were prioritized as candidates. Abbreviations: AD, autosomal dominant; AR, autosomal recessive; CMGG, Center for Medical Genetics Ghent; GE, Genomics England; LCA, Leber Congenital Amaurosis; MAF, minor allele frequency; TSS, transcription start site; uORF, upstream open reading frame.

Four of these variants were found to alter splicing in genes linked to autosomal dominant (*PRPF31*), autosomal recessive (*NMNAT1*) and X-linked (*NDP*) disease. The *NDP*:c.-70G>A and *PRPF31*:c.-9+1G>A variants have recently been reported by Daich Varela *et al*. (2022)^48^. We identified a *de novo* variant (*PRPF31*:c.-9+1G>T) in the same 5’UTR position of *PRPF31* in a sporadic case with rod-cone dystrophy in which bi-allelic *GRM6* variants had been identified, one of them reported as likely benign (**Table S14**). Variants in *GRM6* cause autosomal recessive congenital stationary night blindness^89^, which is different from the clinical diagnosis in this proband. The *NMNAT1:*c.-57G>A variant was found in an individual with macular dystrophy that carries *in trans* an extremely rare missense variant for which *in silico* predictions support a pathogenic effect.

Changes in secondary structure were predicted for 4 variants in genes linked to autosomal dominant (*ARL3, PAX6*) and autosomal recessive (*MERTK, RD3*) disease (**Figure S6**). Given the inheritance pattern and *in silico* predictions, a gain-of-function effect was hypothesized for the *ARL3*:c.-88G>A and *PAX6*:c.-44T>C variants. Both the *MERTK*:c.-125G>A and *RD3*:c.-394G>A variants were identified in homozygous state and found to overlap their respective TSS.

The introduction of an uORF was predicted for 2 variants in the *NPHP4* and *RDH12* genes. In contrast to the uORF created by the *RDH12*:c.-123C>T variant, the one introduced by the *NPHP4*:c.-21C>T variant is out-of-frame and overlaps the CDS. Although no segregation could be established for this case with non-syndromic rod-cone dystrophy, an ultra-rare missense *NPHP4* variant with pathogenic *in silico* predictions was identified.

Only one variant in a gene linked to dominant disease (*PRPF4*) was predicted to affect the primary Kozak consensus sequence. The *PRPF4*:c.-6C>T variant results in a transition to an infrequent nucleotide at this position within the Kozak consensus sequences derived from the IRD gene isoforms selected in this study (**Figure S7**). This variant was found in a sporadic case with rod-cone dystrophy in which 2 likely pathogenic variants have been identified in the *ITF140* gene (**Table S14**); segregation analysis was not possible to establish the phase of any of these alleles.

Four of the variants (*ARL3*:c.-88G>A, *MERTK*:c.-125G>A, *PAX6*:c.-44T>C, *RDH12*:c.-123C>T) were selected for functional validation using diverse multiple experimental approaches.

### Downstream functional analyses support the pathogenicity of the *MERTK*:c.-125G>A and *RDH12*:c.-123C>T variants

Depending on the tissue-specific gene expression and availability of patient material, we conducted either *in vitro* evaluation in ARPE-19 cells (dual luciferase assays or overexpression), or mRNA expression analysis in patient-derived lymphocyte cultures (**Figure 4A**). The *RDH12*:c.-123C>T and *MERTK*:c.-125G>A variants were found to result in a significant decrease in luciferase activity (**Figure 4B**). To further elucidate the underlying mechanism, *Renilla* mRNA expression analysis was performed. Relative *Renilla* luciferase mRNA levels remain unchanged for *RDH12*:c.-123C>T, whereas a significant decrease was observed for *MERTK*:c.-125G>A (**Figure 4C**), hence suggesting a translational and a transcriptional effect for these variants, respectively.

**Figure 4.**
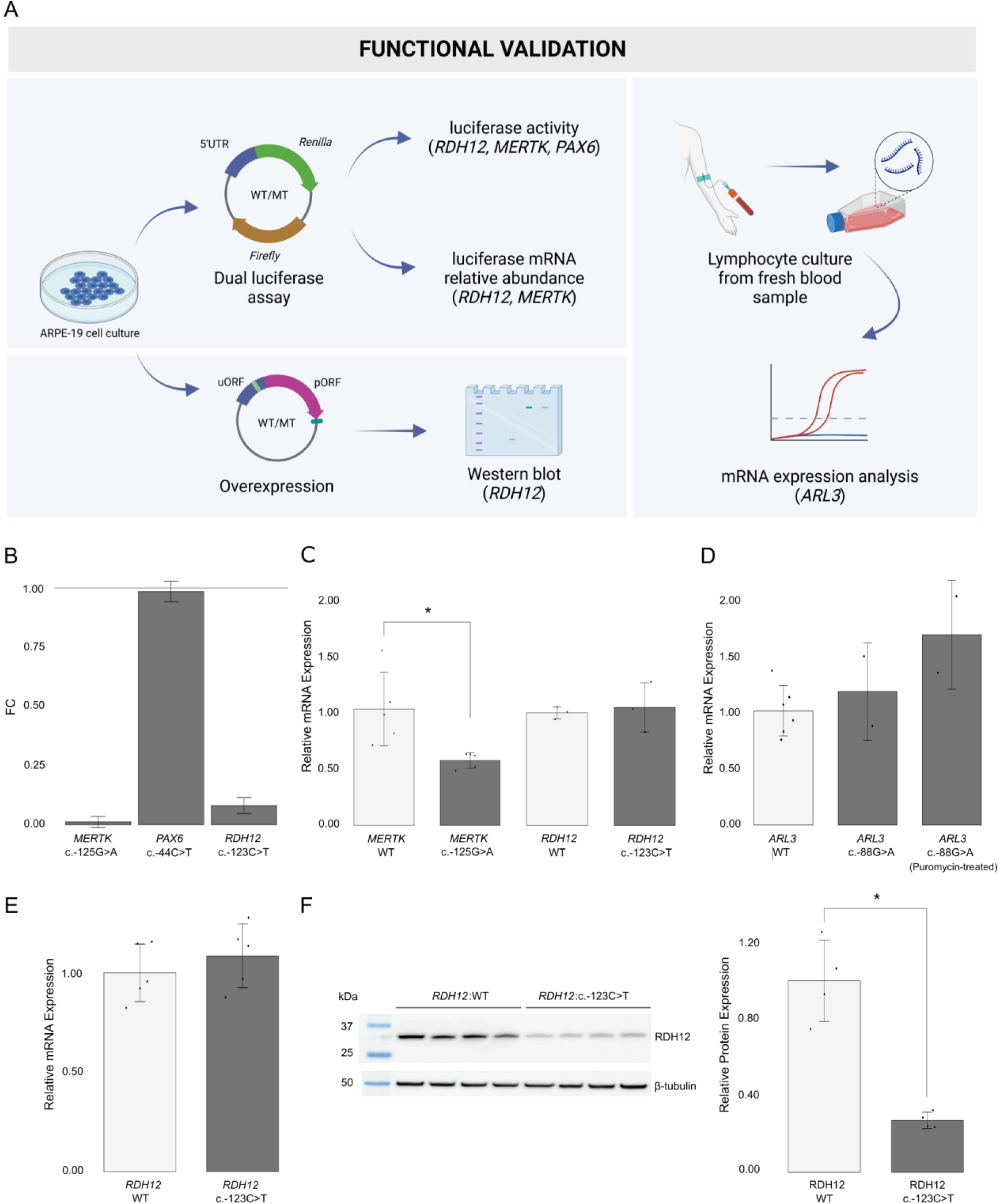
Functional evaluation of candidate pathogenic 5’UTR variants in the *ARL3*, *MERTK*, *PAX6*, and *RDH12* genes. **(A)** Various approaches were used for functionally evaluating candidate variants, including *in vitro* studies (dual luciferase reporter assays and overexpression) and experiments with clinically-accessible tissues (expression analysis in patient-derived lymphocytes). **(B)** Results from the luciferase assays for the *MERTK*:c.-125G>A, *PAX6*:c.-44C>T, and *RDH12*:c.-123C>T variants. The bar plot shows, for each variant, the fold change (FC) of the luciferase reporter level relative to the level of their corresponding wild-type (WT) construct luciferase vector (FC= 1). The *RDH12*:c.-123C>T and *MERTK*: c.-125G>A variants resulted in significant (p<0.001) decrease in luciferase activity (~92% and ~99%, respectively). **(C)** Relative *Renilla* luciferase mRNA levels were significantly decreased (~42%, p<0.01) for the *MERTK*:c.-125G>A variant while they remain the same for the *RDH12*:c.-123C>T variant when normalized to mRNA of *Firefly* luciferase and compared to their corresponding wild-type (WT) construct luciferase vectors. **(D)** qPCR quantification of *ARL3* mRNA abundance in lymphocyte cDNA of two affected siblings carrying the *ARL3*:c.-88G>A novel variant and five healthy controls. No significant differences were observed in *ARL3* mRNA abundance between the affected carriers and controls. **(E-F)** Using an overexpression setting, the *RDH12*:c.-123C>T variant was shown to result in (**E**) unaltered mRNA levels but (**F**) significantly reduced (~73%, p<0.01) RDH12 protein levels.

Taking advantage of the expression of *ARL3* in accessible tissues, we performed qPCR-based quantification of *ARL3* mRNA abundance in lymphocyte cDNA from the two affected siblings in whom we identified the *ARL3*:c.-88G>A variant and five healthy controls. No significant differences were observed in *ARL3* mRNA abundance between the affected carriers and controls (**Figure 4D**). Interestingly, *ARL3* expression levels were slightly higher in the patient samples derived from the lymphocyte cultures treated with puromycin, a translation inhibitor used to suppress nonsense-mediated mRNA decay, compared to the corresponding untreated samples and controls. Therefore, although the *ARL3*:c.-88G>A variant was not predicted to affect splicing, we also performed Sanger sequencing on the cDNA derived from the puromycin-treated and untreated patient lymphocyte cultures. Neither splicing defects nor allele-specific expression were observed (*data not shown*).

In view of the negative results obtained for the *PAX6*:c.-44T>C and *ARL3*:c.-88G>A variants, the phenotypes of patients F9 and F10 were re-evaluated. Neither anterior segment abnormalities nor other clinical presentations compatible with a *PAX6*-related disease were observed in patient F9 or her affected relatives. Regarding F10, re-evaluation of one of the affected siblings revealed an acquired vascular ocular condition instead of an IRD.

### The *RDH12*:c.-123C>T 5’UTR variant is found always *in cis* with the p.Arg234His hypomorphic allele and results in reduced RDH12 protein levels

We identified the *RDH12*:c.-123C>T variant in one solved case from the GE cohort characterized by bi-allelic *RDH12* coding variants (c.[701G>A];[c.735_743del], p.[Arg234His]; [Cys245_Leu247del]). The 5’UTR variant, reported as VUS in ClinVar, was found in *cis* with the p.Arg234His allele. Further evaluation of this 5’UTR variant in 10 additional *RDH12* bi-allelic patients from 8 families carrying the p.Arg234His variant revealed that the c.-123C>T and p.Arg234His variants always form a complex allele (**Table S15**). This was also shown for 7 carriers of the p.Arg234His variant affected by other pathologies (*data not shown*).

Here, we report the first patient who is homozygous for the *RDH12*: c.[-123C>T];[701G>A] complex allele (**Table S15**). Compared to other patients carrying the 5’UTR variant *in trans* with a null *RDH12* allele^90^, patient F21 presents with a milder phenotype (mild decreased acuity (20/25 OD and OS) in fourth decade of life, foveal sparing maculopathy with a circumscribed area of atrophy within the vascular arcades, without a nasal component.

The c.-123C>T variant is predicted to introduce an uAUG into a strong Kozak consensus sequence that is in-frame with a stop codon located 75 nucleotides downstream in the 5’UTR. In view of the results obtained in the luciferase assays and the predicted uORF-introducing effect of the 5’UTR variant, we further inspected the potentially exclusive effect at the translational level by assessing RDH12 mRNA abundance and protein levels in an overexpression setting. Although we could not confirm translation of the 25-amino-acid-long peptide predicted to be encoded by the uORF by co-immunoprecipitation (*data not shown*), we observed unaltered mRNA but significantly (p<0.05) reduced RDH12 protein levels, thereby providing further evidence for a post-transcriptional or translational effect (**Figure 4E-F**) which was already suggested by the mRNA evaluation in the luciferase assays.

## Discussion

Given the emerging role of non-coding variation underlying IRDs^45,48, 52–55^ and the essential regulatory function of 5’UTRs^2,6^, we set out to evaluate their contribution to this heterogeneous group of disorders by analyzing, systematically annotating, filtering and prioritizing 5’UTR variants in two large IRD cohorts in combination with different experimental approaches for functional validation.

Thus far, only a few studies have implicated 5’UTR variation in the molecular pathogenesis of IRD cases^48,53,54^. Our retrospective analysis of 5’UTR variants in IRD genes listed in the ClinVar database^66^ revealed that as many as 58% of all variants have been classified as VUS. A large fraction of the underrepresentation of (likely) pathogenic 5’UTR variants in clinical databases can be explained by their exclusion from downstream tiering pipelines, which prioritize variants with protein-altering consequences and hence neglect the potential impact of 5’UTR variants^91^. Furthermore, the identification of these variants is not always feasible by means of commercial exome enrichment platforms, in particular when variants are found within 5’UTR exons that are not part of or proximal to the first protein-coding exon^19^. Here, we show that the enriched 5’UTR fraction is highly dependent on the WES capture kit used, and ranges from 7% to 39%. Overall, as previous studies have also argued^92–94^, our analysis underscores the diagnostic value of re-analyzing exome data, in which causative variants might already be present but previously disregarded.

Furthermore, the identification of the genetic defects underlying IRDs is greatly impacted by the inherent complexity of the retinal transcriptional landscape. Variants in retina-enriched isoforms and tissue-specific mis-splicing have been shown to be important molecular mechanisms underpinning disease pathogenesis and phenotypic heterogeneity^95–99^. As gene isoforms can display differential 5’UTRs that can result in differential translational efficiencies^100^, we performed a transcript-level re-analysis of retinal expression data to obtain a relevant selection of 5’UTRs of IRD genes for downstream variant analysis. We identified 76 IRD genes with alternative isoforms exhibiting retinal expression levels higher than their respective canonical isoforms, of which 20 displayed a fully distinct 5’UTR. This analysis also found the recently identified photoreceptor-specific non-canonical *CRB1* isoform, which bears a unique 5’UTR exon^101^. Similarly, we confirmed the retinal enrichment of an alternative *RIMS2* isoform containing an unconventional 5’UTR exon, which has been shown to be photoreceptor-specific and functionally conserved in mouse (Del Pozo-Valero *et al*., *unpublished data*). By revealing that a significant fraction of IRD genes express alternative retinal isoforms, our analysis highlights the importance of isoform-aware variant annotation for adequate interpretation.

To aid the assessment of 5’UTR variant pathogenicity, *in silico* tools have recently been developed, with an emphasis put on uORF-perturbing variants^20,26^. A systematic characterization of this class of 5’UTR variants showed that they are subject to strong negative selection, which could even be equivalent to that observed against missense variants^11^. However, thus far, these tools do not provide comprehensive annotations of all possible effects 5’UTR variants can exert^8^. Here, we designed an integrative annotation of 5’UTR-informative features and built a prioritization strategy combining family and phenotypic data to increase the probability of identifying potential pathogenic 5’UTR variants. Although we performed these analyses in IRD cohorts, the features evaluated, and the methods presented in this study can be extrapolated and applied in any setting involving clinical interpretation of 5’UTR variation.

We prioritized 11 candidate pathogenic 5’UTR variants. The great majority of these variants (8/11) have not been reported before, most likely due to their exclusion from routine prioritization pipelines and subsequent clinical interpretation^91^. These 11 variants were assigned to 5 of the 7 categories we defined to classify their potential functional consequences, thus reflecting the diversity in predicted functional effect. Of note, the majority (4/11) was predicted to have an effect on splicing which, in line with our analysis revealing that almost one third of all IRD genes have their 5’UTRs spliced, supports that these regions are also susceptible to disease-causing splicing defects. Notably, we also provided a set of filtered annotated 5’UTR variants with *in silico* predictions pointing to a pathogenic effect that were not prioritized as candidates due to inconsistent genotypes or phenotypes in the patients evaluated. Therefore, we cannot dismiss the possibility that these variants can have a pathogenic effect when present in the right context, e.g. *in trans* with a second pathogenic allele in a gene linked to recessive disease.

Given the particular importance of providing functional evidence to support the pathogenicity of variants located within non-coding regions, we evaluated the predicted functional consequence of 4 candidate variants. Despite the extreme rarity and disease co-segregation of the *PAX6*:c.-44C>T variant, both the lack of functional evidence and the unusual phenotype established upon clinical re-evaluation indicate that this variant is not explaining the phenotype. Likewise, we could not confirm the predicted effect of the *ARL3*:c.-88G>A variant using accessible patient-derived material. The sibling of the proband was eventually considered not affected by an IRD, indicating that the variant is either not causative, incompletely penetrant, or simply a recessive allele. Interestingly, a phenotype similar to that of the proband has been associated with a pathogenic variant in this gene^102^, which could suggest that this variant exerts a retina-specific effect that cannot be evaluated with the current experimental setting. Nevertheless, it is important to note that in this case both siblings have been studied by WES, but were not tested for known deep-intronic and structural variants. Before proceeding with further functional characterization of the *ARL3*:c.-88G>A variant, WGS should thus be performed to confirm or rule out such variants. Altogether, these results highlight the need and potential limitations of experimental assays to validate the functionality of non-coding variants.

We could provide a novel molecular diagnosis in a patient with severe early-onset retinal dystrophy that remained unsolved after WGS screening. This individual was found to be a homozygous carrier for the extremely rare c.-125G>A variant in *MERTK*, overlapping its transcription start site, for which our *in silico* predictions pointed to an effect at the transcriptional level. A drastic reduction of luciferase activity accompanied by decreased relative *Renilla* luciferase mRNA levels confirmed the hypothesized transcriptional effect of the variant, which is consistent with a loss of function of *MERTK*. To our knowledge, this is the first reported disease-causing 5’UTR variant in *MERTK*.

One of the candidate pathogenic 5’UTR variants was found in a case solved with bi-allelic *RDH12* coding variants, namely the p.Arg234His hypomorphic missense^90,103,104^ and the p.Cys245_Leu247del in-frame variants. The identified c.-123C>T 5’UTR variant was found *in cis* with p.Arg234His, both in this case as well as in 10 additional *RDH12* bi-allelic patients screened for this 5’UTR variant, with one of them being homozygous for this complex allele. The pathogenicity of p.Arg234His has been questioned in view of functional assays revealing RDH12 protein levels and catalytic activity comparable to the wild-type or polymorphic alleles, respectively^105^. We therefore hypothesized a pathogenic effect exerted by the *RDH12*:c.-123C>T variant as part of this complex allele. This 5’UTR variant was predicted to introduce an upstream start codon (uAUG) into a strong Kozak consensus, thereby creating an uORF. Given that ribosomes would first encounter and start translating from this gained uAUG, we hypothesized that this variant would decrease normal RDH12 translation and thus result in lower protein levels. Furthermore, this variant has been argued to affect the function of an alternative promoter of *RDH12* based on the observed decreased activity in a luciferase reporter assay^52^. Importantly, this experimental setup did not include comparison between mRNA levels and luciferase protein activity and was therefore unable to pinpoint at which level the effect occurs. Here, using a dual luciferase assay with the 5’UTR instead of the genomic sequence, followed by downstream evaluation at both the mRNA and protein level, we confirmed the hypothesized uORF-mediated effect. We tested this hypothesis further using an overexpression setting and demonstrated unaltered mRNA but significantly reduced RDH12 protein levels, hence confirming the translational effect of *RDH12*:c.-123C>T variant. Although we could not obtain experimental evidence supporting the translation of the uORF-encoded peptide, which might suggest its instability, we cannot rule out alternative mechanisms such as ribosome stalling^9^. However, further studies are needed to confirm this hypothesis. Overall, our results indicate that not p.Arg234His alone, but rather the combination of p.Arg234His and c.-123C>T, or even c.-123C>T by itself, is disease-causing (**Figure 5**).

**Figure 5.**
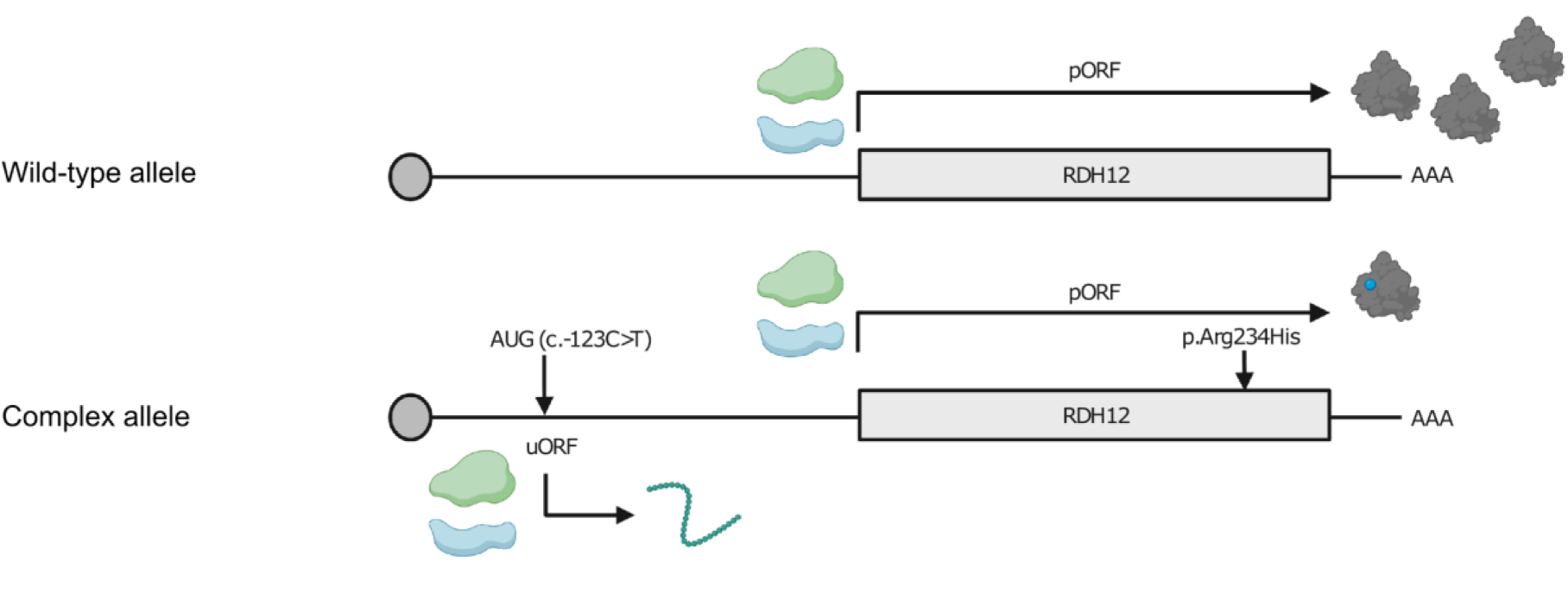
Proposed pathogenetic mechanism of the *RDH12*: c.[-123C>T];[701G>A] complex allele. Graphical representation of the *RDH12* wild-type allele (*top*) and the *RDH12* complex allele harboring the c.701G>A (p.Arg234His) and the c.-123C>T variants (*bottom*). The 5’UTR variant introduces an upstream start codon into a strong Kozak sequence which can be recognized by ribosomes. Either initiation or active translation of the introduced upstream open reading frame (uORF) results in lower translational efficiency of the primary open reading frame (pORF). As a result, there is a decreased level of RDH12 protein with the arginine-to-histidine amino acid substitution at position 734 (*blue point*).

## Conclusions

5’UTRs are essential modulators of post-transcriptional and translational control. Even though they can be captured by WES to a large extent, variants within these regions are typically overlooked in both research and routine clinical settings, mainly due to their challenging interpretation. In this study, we developed a systematic strategy to comprehensively annotate and prioritize 5’UTR variants in IRD genes. This strategy combined with functional studies allowed us to pinpoint 5’UTR variants providing either novel molecular diagnoses or the full picture explaining the pathogenicity of previously reported hypomorphic variants. Overall, in this study we highlight the importance of multi-layered annotation and validation of non-coding variation potentially underlying disease and provide a 5’UTR interpretation approach that could be extrapolated to other rare diseases.

## Data Availability

The data that support the findings of this study are available within the Genomics England (protected) Research Environment but restrictions apply to the availability of these data, as access to the Research Environment is limited to protect the privacy and confidentiality of participants. Likewise, healthcare and genomic data derived from individuals included in the Center for Medical Genetics Ghent (CMGG) cohort are not publicly available to comply with the consent given by those participants. De-identified data as well as analysis scripts are available from the authors upon reasonable request. Extended data generated in this study are available in the supplementary materials.

## Abbreviations

5’UTR: 5’ untranslated region
AD: Autosomal dominant
AR: Autosomal recessive
CAGE-seq: Cap analysis gene expression sequencing
CMGG: Center for Medical Genetics Ghent
CDS: Coding sequence
DMEM: Dulbecco’s minimal essential medium
ERDC: European Retinal Disease Consortium
FC: Fold change
GE: Genomics England
IRD: Inherited retinal disease
IRES: Internal ribosomal entry sites
MAF: Minor allele frequency
OD/OS: *Oculus dexter*/*sinister* (right/left eye)
qPCR: Quantitative polymerase chain reaction
SNV: Single-nucleotide variant
SV: Structural variant
TSS: Transcription start site
TPM: Transcripts per million
TE: Translational efficiency
uORF: Upstream open reading frame
VUS: Variant of uncertain significance
WES/WGS: Whole exome/genome sequencing

---

**Figure S1.**
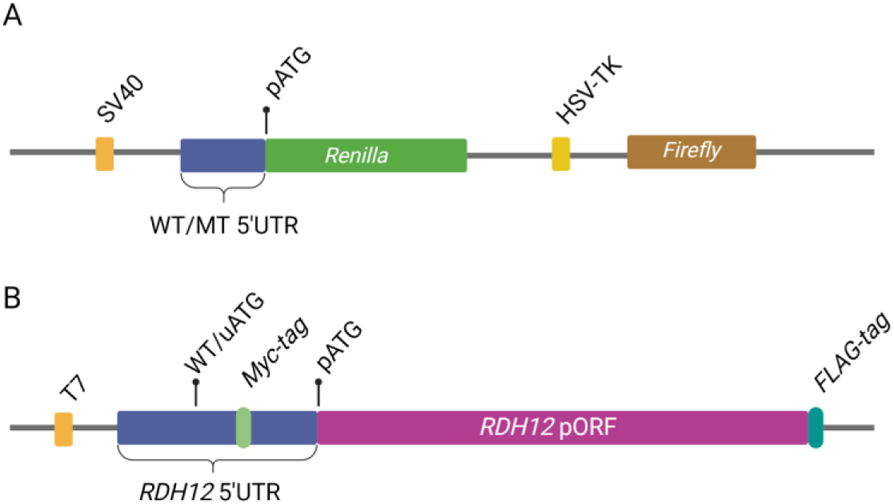
Schematic overview of constructs used for functional studies. (**A**) The wild-type (WT) 5’UTRs of the *MERTK*, *PAX6*, and *RDH12* genes were cloned in-frame into a psiCHECK™-2 dual luciferase vector (Promega), containing the *Renilla* and *Firefly* luciferase reporter genes under the regulation of the SV40 and HSV-TK promoters, respectively. (**B**). Depiction of the overexpression RDH12 construct comprising the 5’UTR and primary open reading frame (pORF) of *RDH12*, for which Myc and FLAG in-frame tags were included downstream, respectively. This fragment was cloned into a pcDNA™3.1^(+)^ (Invitrogen) vector downstream its T7 promoter. For all constructs, 5’UTRs variants were created by site-directed mutagenesis to obtain mutant (MT) constructs with the variants of interest.

**Figure S2.**
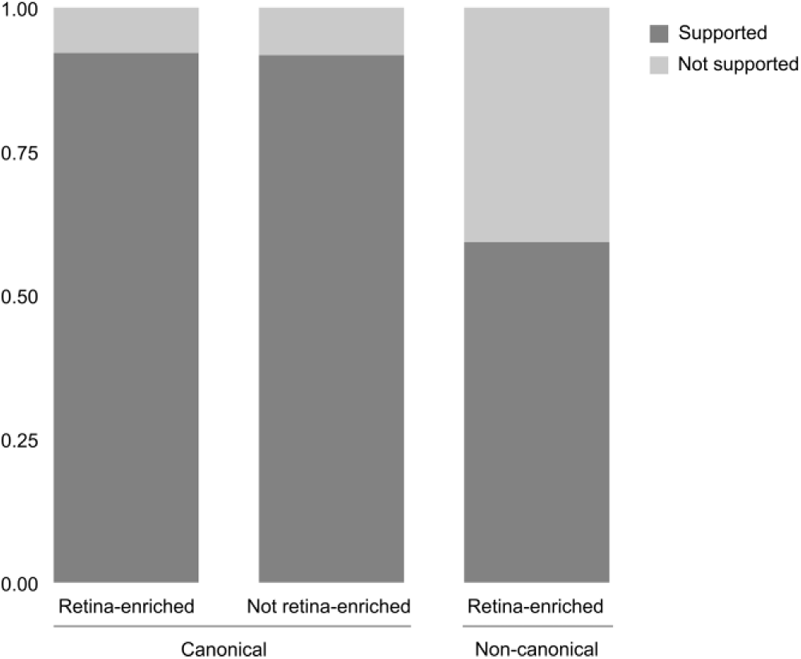
Assessment of transcription start site (TSS) confidence by CAGE-seq in retina. Representation of the proportion of IRD gene isoforms based on the support of their annotated TSS provided by CAGE-seq data derived from adult and fetal retina.

**Figure S3.**
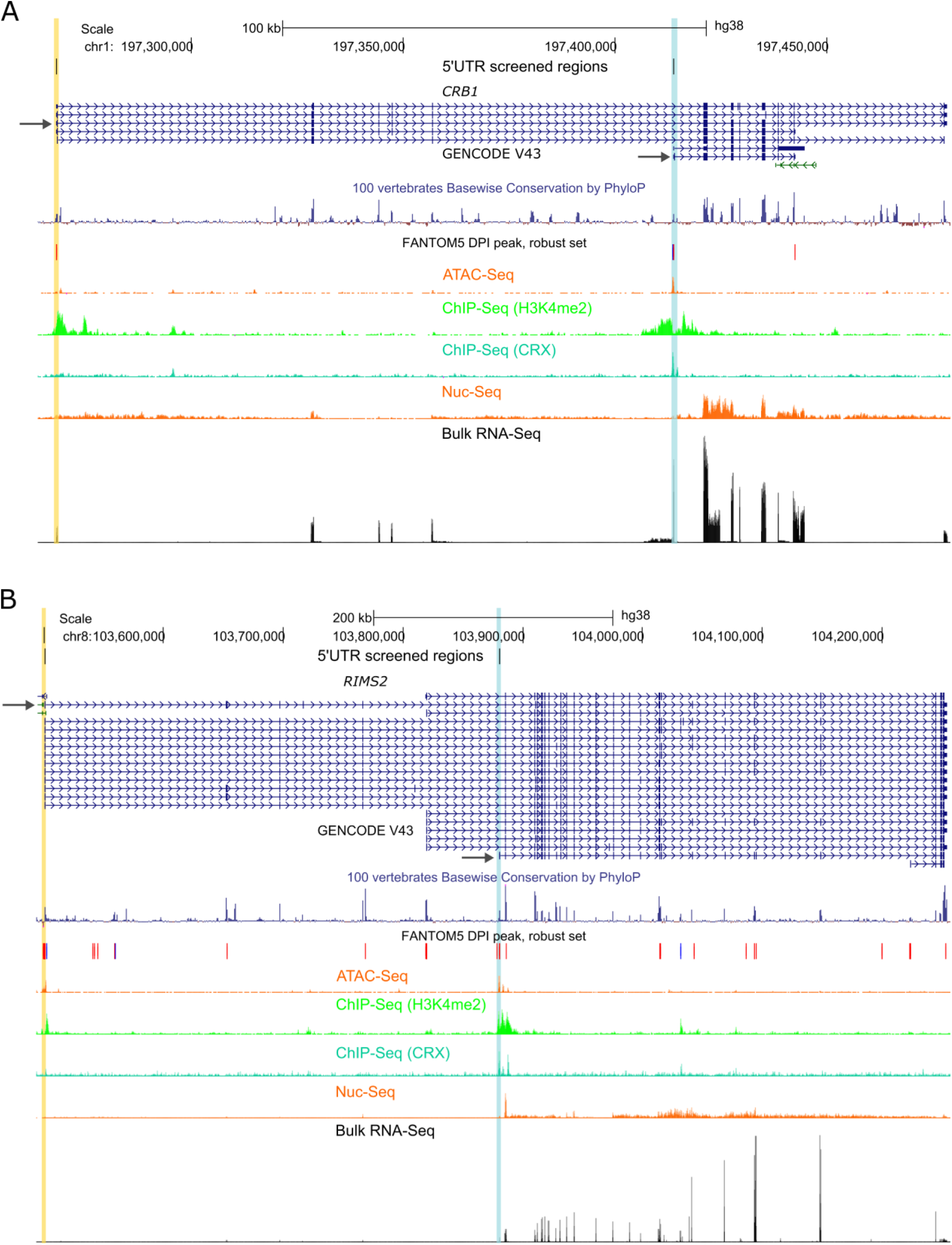
Retina-enriched non-canonical isoforms of the *CRB1* and *RIMS2* genes. The shorter alternative isoforms of (**A**) *CRB1* (ENST00000681519) and (**B**) *RIMS2* (ENST00000436393) contain 5’UTRs which are completely distinct to those of their respective canonical isoforms (ENST00000367400.8 and ENST00000696799.1, respectively). The transcription start sites (supported by CAGE-seq) of each canonical and non-canonical isoform (indicated by arrows) are highlighted in *yellow* and *blue*, respectively. Active retinal transcription is supported by bulk RNA-seq and Nuc-seq derived from human retina. The enrichment in retina of these isoforms is further supported by signatures of open chromatin, H3K4me2, and binding of the retina-specific transcription factor CRX (datasets used are listed in **Table S2**).

**Figure S4.**
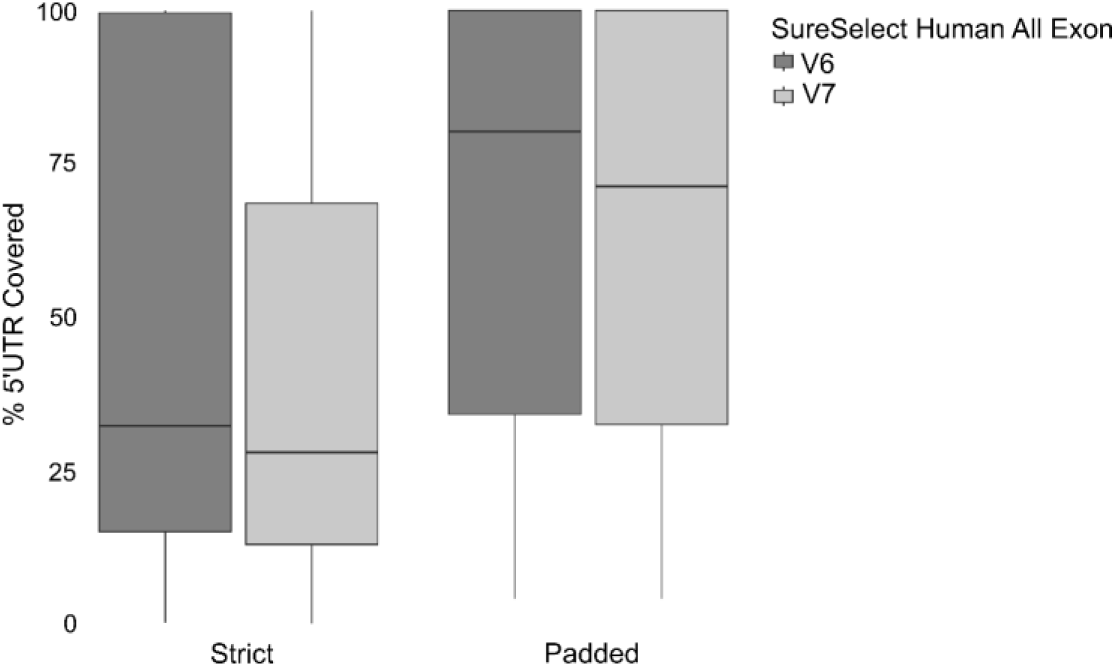
Evaluation of 5’UTR capture by the kits mostly used for generating our in-house WES data. Comparison of the capture performance of the SureSelect Human All Exon V6 and V7 kits (Agilent Technologies) considering *strict* or *padded* designs (*see Methods*).

**Figure S5.**
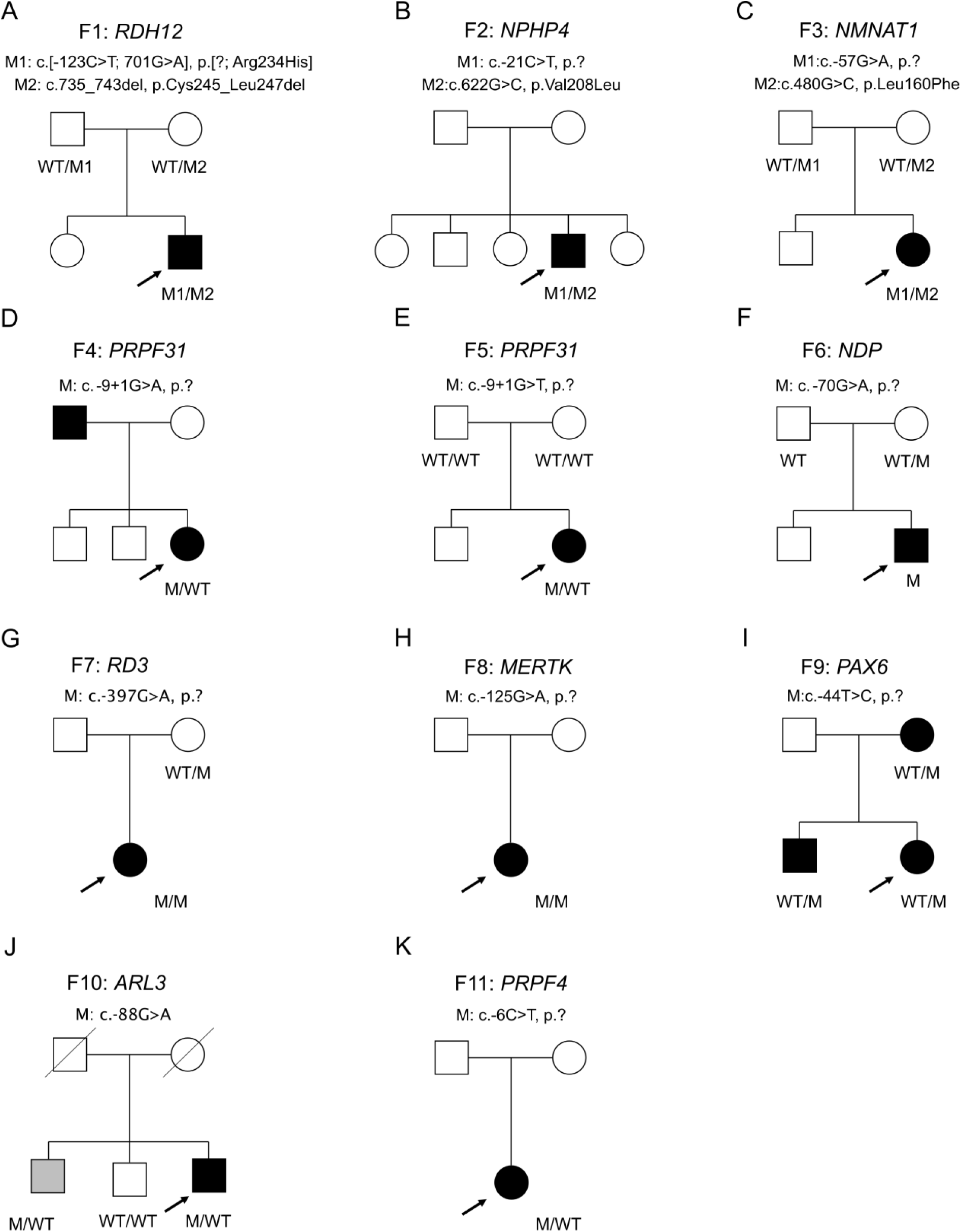
Pedigrees of families carrying the 11 candidate 5’UTR variants and segregation analysis. (**J**) Upon clinical re-evaluation, the sibling of F10 (*gray*) was found to be affected by an acquired vascular ocular condition instead of an IRD. Abbreviations: WT, wild-type allele; M, mutated allele.

**Figure S6.**
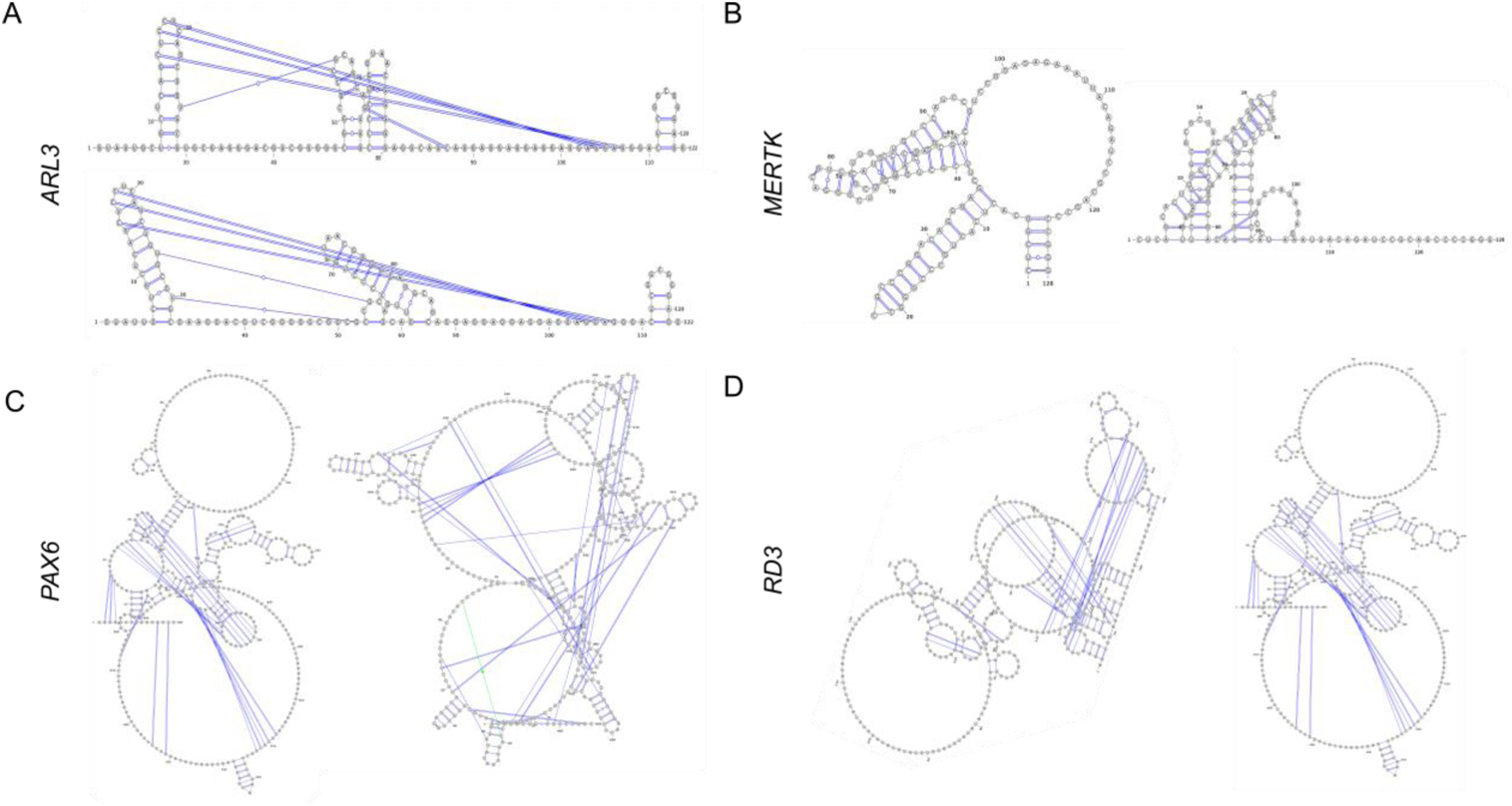
Secondary structure prediction for wild-type and mutated 5’UTR sequences. Secondary structure analysis of the 5’UTR sequences (*Ufold* using default parameters) revealed different folding for the **(A)** *ARL3*:c.-88G>A, **(B)** *MERTK*:c.-125G>A, **(C)** *PAX6*:c.-44T>C, **(D)** *RD3*:c.-394G>A variants in comparison to their corresponding wild-type 5’UTRs.

**Figure S7.**
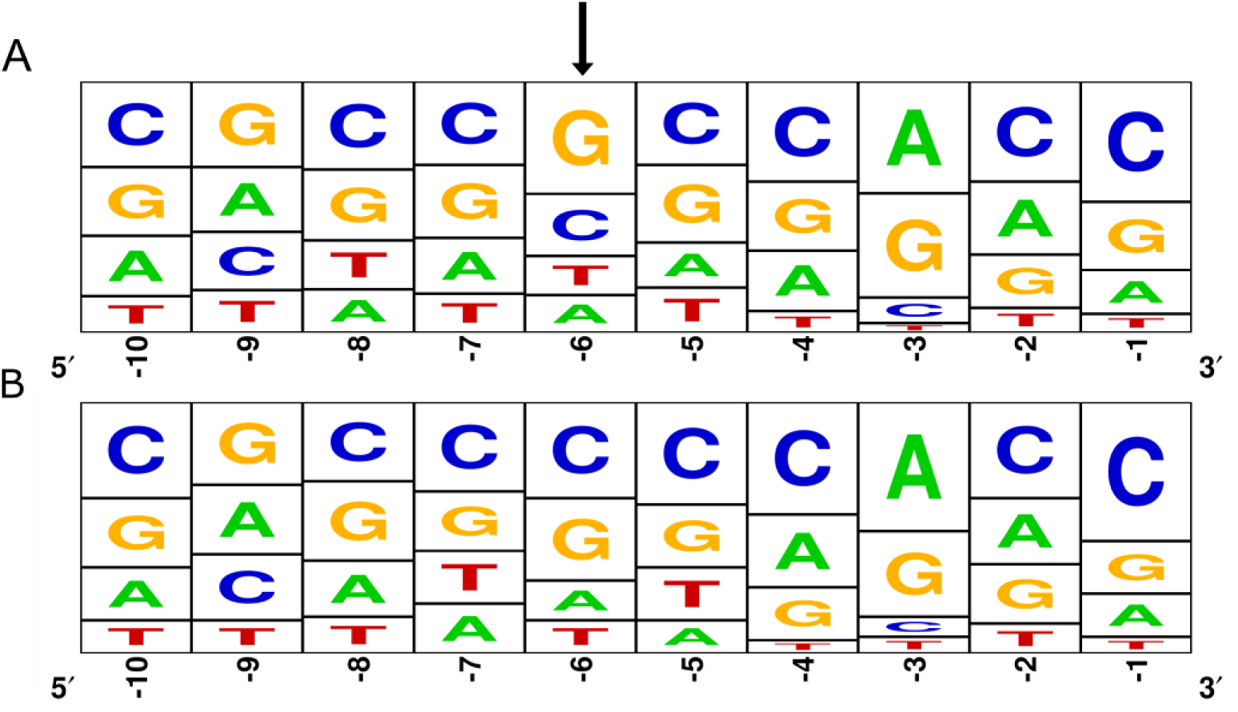
Kozak consensus sequence of all IRD gene isoforms evaluated in this study. The nucleotides within the −1 to −10 positions relative to the main AUG were retrieved for all selected transcripts. Frequency plots corresponding to the Kozak sequences of the (**A**) retina-enriched (373) and (**B**) not retina-enriched canonical transcripts (76).

## Additional files

### Additional file 1 - Supplementary Tables

**Table S1.** Custom diagnostic gene panel comprising all IRD genes listed in either the Retinal disorders panel (v2.195) from Genomics England PanelApp or RetNet. Abbreviations: AD, autosomal dominant; AR, autosomal recessive; HGNC, HUGO Gene Nomenclature Committee.

**Table S2**. Publicly available multi-omics datasets derived from human retina used in this study.

**Table S3**. Overview of the IRD sub-cohort from the Rare Disease arm of the 100,000 Genomes Project (Genomics England).

**Table S4**. Primer sequences used in this study.

**Table S5.** Overview of selected canonical and non-canonical protein-coding transcripts of IRD genes and their expression in adult human retina. Abbreviations: MANE, Matched Annotation from NCBI and EMBL-EBI; SD, standard deviation; TPM, transcripts per million; TSS, transcription start site.

**Table S6**. Genomic coordinates (GRCh38) of the regions in which variants were searched in this study (*5’UTR analysis file*).

**Table S7**. Overview of the structure of 5’UTRs of the transcripts included in this study.

**Table S8**. Comparison of 5’UTRs of non-canonical transcripts with respect to their corresponding canonical isoforms.

**Table S9**. Overview of the performance of commercial exome capture designs on the 5’UTRs screened in this study.

**Table S10**. Overview of the performance of commercial exome capture kits mostly used for the generation of our in-house WES data considering strict and padded designs.

**Table S11**. Overview of all 5’UTR variants reported in IRD genes submitted to the ClinVar database.

**Table S12.** Overview of the span of internal ribosomal entry sites (IRES) contained in the 5’UTR of IRD genes.

**Table S13**. Illustration of the designed prioritization strategy applied to a selection of the identified variants. Variants which did not pass all criteria (*strikethrough*) were not prioritized as candidates (*bold*).

**Table S14**. Overview of additional variants reported for patients F5, F8, F11. Abbreviations: ACMG, American College of Medical Genetics and Genomics; MAF, minor allele frequency. **Table S15**. Overview of the 11 patients carrying the *RDH12*:c.701G>A (p.Arg234His) variant found *in cis* with *RDH12*:c.-123C>T. Abbreviations: ARG: Argentina; CMGG, Center for Medical Genetics Ghent; GE, Genomics England; HU FJD: Hospital Universitario Fundación Jiménez Díaz; IOB: Institute of Molecular and Clinical Ophthalmology Basel; MEH: Moorfields Eye Hospital.

### Additional file 2 - Dataset S1

Annotation of the 1,450 5’UTRs variants that remained after functional classification and filtering. For each cohort (GE or CMGG) there is one tab per functional category.

## Declarations

### Ethics approval and consent to participate

The 100,000 Genomes Project Protocol has ethical approval from the HRA Committee East of England – Cambridge South (REC Ref 14/EE/1112). This study was registered with Genomics England within the *Hearing and sight domain* under Research Registry Projects 465. This study was approved by the ethics committee for Ghent University Hospital (B6702021000312) and performed in accordance with the tenets of the Helsinki Declaration and subsequent reviews.

### Consent for publication

Not applicable. We present only de-identified data.

### Authors’ contributions

ADR: Conception and project design, acquisition of data, analysis and interpretation of data, drafting and revising the manuscript.

MDPV: Acquisition of data, analysis and interpretation of data, drafting and revising the manuscript.

MB: Acquisition of data, analysis and interpretation of data, revising the manuscript.

FVB: Acquisition of data, revising the manuscript.

MDV: Acquisition of data, revising the manuscript.

MVH: Acquisition of data, revising the manuscript.

MDB: Acquisition of data, revising the manuscript.

SVS: Acquisition of data, revising the manuscript.

MBW: Acquisition of data, revising the manuscript.

JE: Acquisition of data, revising the manuscript.

GER: Acquisition of data, revising the manuscript.

CR: Acquisition of data, revising the manuscript.

AW: Acquisition of data, revising the manuscript.

GA: Acquisition of data, revising the manuscript.

CA: Acquisition of data, revising the manuscript.

JDZ: Acquisition of data, revising the manuscript.

BL: Acquisition of data, revising the manuscript.

EDB: Project supervision, acquisition of data, revising the manuscript.

FC: Conception and project supervision, acquisition of data, analysis and interpretation of data, revising the manuscript.

## Acknowledgements

This research was made possible through access to the data and findings generated by the 100,000 Genomes Project. The 100,000 Genomes Project is managed by Genomics England Limited (a wholly owned company of the Department of Health and Social Care). The 100,000 Genomes Project is funded by the National Institute for Health Research and NHS England. The Wellcome Trust, Cancer Research UK and the Medical Research Council have also funded research infrastructure. The 100,000 Genomes Project uses data provided by patients and collected by the National Health Service as part of their care and support. We thank Olga Zurita for their excellent technical assistance. We are grateful to Dr. Jacqueline Cook and Dr. Savita Madhusudhan for their clinical input.

